# Achieving behavior change at scale: Causal evidence from a national lifestyle intervention program for pre-diabetes in the UK

**DOI:** 10.1101/2023.06.08.23291126

**Authors:** Julia M. Lemp, Christian Bommer, Min Xie, Anant Jani, Justine I. Davies, Till Bärnighausen, Sebastian Vollmer, Pascal Geldsetzer

**Affiliations:** Heidelberg Institute of Global Health, Heidelberg University Hospital; Heidelberg, Germany; Department of Economics and Centre for Modern Indian Studies, University of Göttingen; Göttingen, Germany; Oxford Martin School, Oxford University; Oxford, UK; Institute for Applied Health Research, University of Birmingham; Birmingham, UK; Centre for Global Surgery, Department of Global Health, Stellenbosch University; Cape Town, South Africa; Medical Research Council/Wits University Rural Public Health and Health Transitions Research Unit, Faculty of Health Sciences, University of the Witwatersrand; Johannesburg, South Africa; Africa Health Research Institute; Somkhele, South Africa; Department of Global Health and Population, Harvard T.H. Chan School of Public Health, Harvard University; Boston, MA, USA; Division of Primary Care and Population Health, Stanford University; Stanford, CA, USA; Chan Zuckerberg Biohub; San Francisco, CA, USA

## Abstract

There remains widespread doubt among clinicians that mere lifestyle advice and counseling provided in routine care can achieve improvements in health. We aimed to determine the health effects of the largest behavior change program for pre-diabetes globally (the English Diabetes Prevention Programme) when implemented at scale in routine care. We exploited the threshold in glycated hemoglobin (HbA1c) used to decide on program eligibility by applying a regression discontinuity design, one of the most credible quasi-experimental strategies for causal inference, to electronic health data from approximately one-fifth of all primary care practices in England. Program referral led to significant improvements in patients’ HbA1c and body mass index. This analysis provides causal, rather than associational, evidence that lifestyle advice and counseling implemented in a national health system can achieve important health improvements.

---

By 2030, the number of adults with diabetes globally is expected to reach 578 million, representing 10.2% of the global adult population (*1*). Diabetes is an important cause of mortality, morbidity, and health-system costs (*2*). There is, thus, an urgent need to implement population-based interventions that prevent diabetes, enhance its early detection, and address cardiovascular risk factors to prevent or delay its progression to complications. In particular, type 2 diabetes (T2DM), which accounts for approximately 90% of the total diabetes burden, is a major risk factor for cardiovascular disease, with people with diabetes having a more than twofold increase in the risk of incident chronic heart disease compared to those without diabetes (*3*). Worryingly, and at least partly driven by the increase in excess weight and obesity, diabetes prevalence and diabetes-related deaths continue to rise in most parts of the world (*4*).

Often resulting from socioecological contexts, individual behaviors such as poor nutrition, hazardous alcohol consumption, and physical inactivity play a key role in the development of T2DM (*5*). While targeting individual behavior as a preventive strategy for T2DM has been controversial (*6*), behavior change programs (sometimes referred to as lifestyle interventions or similar) have been efficacious in controlled clinical trials (*7, 8*). In the seminal US Diabetes Prevention Program study (which serves as a model for many behavior change programs in the US and elsewhere) (*9, 10*), targeting changes in lifestyle behavior was even more effective than metformin in preventing or delaying diabetes. However, trials such as the US Diabetes Prevention study have mainly focused on efficacy, supplying proof of principle that the intervention worked when one-to-one sessions with specialists and a range of incentives are being provided (*11*). Thus, while a recent meta-analysis concluded that lifestyle modification provides strong evidence for reversing pre-diabetes in adults (*12*), it remains important to establish the transferability of behavior change programs into real-world settings.

Establishing that behavior change programs work in routine care is essential for several reasons. First, although lifestyle counseling is the recognized first-line treatment option for people presenting with pre-diabetes and other cardiovascular risk factors, clinicians often revert to prescribing preventive medication due to limited time resources in primary care (*13*), insufficient knowledge and referral options for promoting healthy lifestyles (*14, 15*), and a predominance of the biomedical model with clinicians being uncertain about the success of counseling (*14, 16*). In particular, doubt that behavior change at the level required for substantial weight loss is possible to achieve for most patients is prevalent among primary care clinicians (*17*). Second, participants enrolling in clinical trials for behavior change programs are unlikely to be representative of the broader patient population. For example, patients enrolled in clinical cardiology trials, compared with patients encountered in everyday practice, had a lower risk profile as they were younger, more likely to be male, and less likely to have a comorbid disease (*18*). Individuals drawn from an unselected, general population may respond differently, given their lower health literacy and willingness to engage, higher comorbidities, and greater ethnic diversity (*19, 20*).

Hence, we advance the argument that the impact of behavior change programs on population health must relate to real-world effectiveness and should, thus, be evaluated in an “observational, non-interventional trial in a naturalistic setting” akin to phase IV in drug development (*21, 22*). However, conventional observational studies that are generally applied in health research have the disadvantage that they may fail to account for major confounding factors such as selection biases and, thus, preclude causal interpretations. In contrast, in this study, we provide causal evidence for the effectiveness of a behavior change program using an innovative causal inference method in large-scale routine data.

We establish causality by applying a regression discontinuity approach that combines a rich set of electronic health records from the English National Health Service with variation in treatment probabilities generated by guidelines from the National Institute for Health and Clinical Excellence (NICE) that recommend intensive lifestyle counseling for people at high risk of progression to T2DM (*23*). Specifically, we exploit the fact that the NHS Diabetes Prevention Programme (NHS DPP), a behavior change program with weight loss, diet, and physical activity goals consisting of at least 13 group sessions over the course of nine months and the largest DPP globally to achieve universal population coverage, is only open to patients above a prespecified threshold of HbA1c or fasting plasma glucose (*24*). This way we can take advantage of existing large-scale routine health data while still obtaining causal effect estimates that are not vulnerable to confounding and measurement error (*25, 26*). Ultimately, our study aims to determine the real-world health effects of the NHS DPP, investigating whether a routine behavior change program in a national health system has the potential to lead to improvements in key cardiovascular risk factors such as HbA1c, excess weight, raised blood pressure, and blood lipid levels.

## Results

Our primary outcome was change in HbA1c. Secondary outcomes included changes in body mass index (BMI), body weight, blood pressure, serum cholesterol levels, and serum triglycerides levels. We also conducted exploratory analyses investigating the effect of program referral on the probability of diabetes, hypertension, and hyperlipidemia incidence; newly prescribed medications for these conditions; diabetic complication; all-cause mortality; and emergency hospitalization for a major adverse cardiovascular event. A detailed definition of each outcome is provided in table S1.

### Data source and sample selection

Our study used data from the Clinical Practice Research Datalink (CPRD) Aurum and NHS England’s Hospital Episode Statistics (HES). CPRD Aurum is a large primary care database of de-identified electronic health records from a network of approximately one-fifth of General Practitioner (GP) practices across England. To ensure sufficient implementation of the NHS DPP during the study period after the start of the phased roll-out in mid-2016, our population of interest consisted of adults (aged 18 to 80 years) who received an HbA1c test between January 1^st^, 2017, and December 31^st^, 2018. Data were available until the end of June 2020. We identified 2 106 376 patients who had a baseline HbA1c test during the enrolment and met inclusion criteria for our primary cohort (see Methods and Materials). 2 052 480 of these (97.4%) had been registered with their GP for at least 6 months following the index date. Patient characteristics are described in Table 1. Their mean age was 51 years, 915 717 (43.5%) were men and 1 190 659 (56.5%) were women, and their mean baseline HbA1c level was 36.6 mmol/mol. The median time to endline HbA1c during follow-up was 20.5 months (interquartile range, 13.5-26.8). Of the 2 052 480 patients who had a follow-up time of at least 6 months, 2 037 384 (99.3%) were linkable to HES hospitalization data. At baseline, 749 884 (36.5%) people had already received at least one prescription for a blood pressure-lowering medication and 411 288 (20.0%) for a lipid-lowering medication.

**Table 1.** Sample characteristics. N = 2 106 376. Abbreviations: BMI, body mass index; GP, General Practitioner. Age, gender, and number of consultations was available for the full sample. Missingness in baseline BMI, blood pressure and lipids are shown in table S2.

| Variable | % | N |
| --- | --- | --- |
| Age in years, mean (SD) | 51 (16) |  |
| 18 to 29 | 11.6 | 243 793 |
| 30 to 39 | 14 | 295 282 |
| 40 to 49 | 19.5 | 411 201 |
| 50 to 59 | 21.9 | 461 006 |
| 60 to 69 | 17.9 | 377 073 |
| 70 to 80 | 15.1 | 318 021 |
| Gender |  |  |
| Female | 56.5 | 915 717 |
| Male | 43.5 | 1 190 659 |
| BMI, mean (SD) | 28.1 (6.1) |  |
| < 25 | 33.1 | 436 041 |
| 25 to 29.9 | 35.2 | 464 217 |
| 30 to 34.9 | 19.6 | 258 266 |
| 35 to 39.9 | 7.7 | 101 321 |
| ≥ 40 | 4.4 | 58 444 |
| Hypertension stage based on systolic and diastolic blood pressure |  |  |
| Normal (< 120 and 80) | 19.3 | 188 906 |
| Prehypertension (120-139 or 80-89) | 59.6 | 583 428 |
| Stage 1 Hypertension (140-159 or 90-99) | 18.6 | 182 074 |
| Stage 2 Hypertension (≥160 or ≥ 100) | 2.5 | 24 222 |
|  | Mean | SD |
| Baseline blood pressure |  |  |
| Diastolic, mmHg | 80 | 11 |
| Systolic, mmHg | 132 | 18 |
| Baseline lipids |  |  |
| HDL, mmol/l | 1.47 | 0.45 |
| LDL, mmol/l | 3.02 | 0.96 |
| Total-cholesterol-to-HDL ratio | 3.77 | 1.25 |
| Triglycerides, mmol/l | 1.55 | 0.97 |
| Number of yearly GP consultations | 12.4 | 10.8 |
| Number of prior emergency hospitalization | 1.13 | 5.37 |

### Meeting the statistical assumptions

In the first part of the analysis, we ensured that all necessary assumptions for a regression discontinuity analysis are met. Importantly, we tested the validity of the continuity assumption (*25*). First, the density distribution of baseline HbA1c should be continuous around the threshold; this would be violated if patients (or providers) could precisely manipulate baseline HbA1c. As shown in fig. S1, there was no indication of heaping or manipulation of HbA1c values around the threshold. Secondly, baseline covariates (such as age or previous contact with health service providers) should be balanced, i.e., continuous, at the threshold. As it is in randomized controlled clinical trials, evidence of balance on baseline observables provides confidence that patients assigned to treatment and control conditions are exchangeable. We showed that this assumption is met by plotting the relationship between baseline HbA1c and potential confounders using third-order global polynomial regressions (Fig. 1). The illustrated balance on baseline observables was statistically supported by local linear regressions that yielded no significant discontinuities at the eligibility threshold (Fig. 1, table S3).

**Fig. 1.**
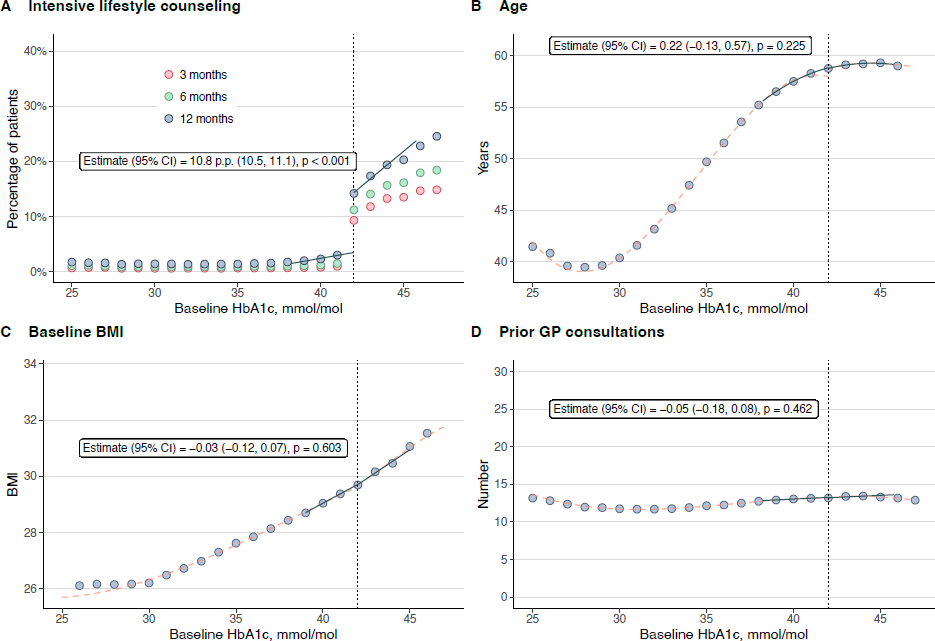
Association between baseline HbA1c and intensive lifestyle counseling and potential confounders. BMI = Body mass index. p.p. = percentage points. (**A**) Primary exposure (‘First stage’) after 3, 6, and 12 months after baseline HbA1c. (**B**), (**C**), and (**D**), Potential confounders. The blue lines show the local linear regression models within the bandwidth used in our primary analysis. The orange dotted lines show the global polynomial relationship. The blue circles represent the mean value for individual patients and the dotted vertical lines indicate the HbA1c cutoff. The estimate represents the discontinuity at the HbA1c threshold, whereas discontinuities in potential confounders may jeopardize assumptions underlying regression discontinuity.

### Program referrals in routine care increase at the eligibility threshold

While increasing HbA1c is associated with an increase in risks of diabetes and cardiovascular disease (*27*), the HbA1c eligibility threshold of 42 mmol/mol (6%) to enter the program does not represent a pathophysiological phenomenon at that specific threshold value. Rather, since the association is continuous and HbA1c was measured with random measurement error, patients just below and above the threshold are close to identical in their underlying characteristics and, thus, effectively randomized to being referred to the program or not. Using a local linear approach, we compared patients lying closely on either side of the threshold which allows for the interpretation of differences in clinical outcomes as causal (*26*).

Local linear regression demonstrated a 10.8 percentage points increase in treatment assignment at the HbA1c eligibility threshold. In relative terms, patients just above the threshold were five times more likely to be referred compared to patients just below the threshold (Fig. 1). Receipt of treatment was defined as a record of a referral to a behavior change program or intensive lifestyle counseling during the 12 months after the baseline HbA1c test. Treatment primarily included referrals to the NHS DPP, but we also included referrals to other structured programs and intensive lifestyle counseling as they are likely to serve as an alternative where placement in NHS DPP is not possible. 26 970 patients with a baseline HbA1c between 42 and 47 mmol/mol were referred to a behavior change program or intensive lifestyle counseling, of which 20 963 (77.7%) were referred to the NHS DPP. 4 800 patients declined NHS DPP referrals offered by their GP. All records considered as treatments are listed in table S4. For convenience only, we henceforth refer to these treatments simply as intensive lifestyle counseling.

### Glycemic control is improved

Patients who were referred to intensive lifestyle counseling significantly improved their HbA1c levels. Specifically, we evaluated the effect of referral to intensive lifestyle counseling on glycemic control by fitting separate regression lines of the association between baseline HbA1c and change in HbA1c above and below the eligibility threshold. The difference in where these lines intersect the threshold quantifies the discontinuity in the outcome and can be described as the intention-to-treat effect of the threshold rule (−0.10 mmol/mol, 95% CI −0.16, −0.03). The intention-to-treat effect measures the effect of being eligible for intensive lifestyle counseling as determined by the guideline rather than the effect of actually being referred and is therefore dependent on the probability of treatment at the threshold, i.e., how many GPs adhere to the guidelines. Thus, to obtain the true effect of being referred to intensive lifestyle counseling (the complier average causal effect), it is necessary to scale the intention-to-treat effect by the difference in the probability of treatment at the threshold. When doing this, we find a significant negative effect of referral to intensive lifestyle counseling on HbA1c at follow-up (−0.85 mmol/mol, 95% CI −1.46, −0.24).

While the clinical significance of a 0.85 mmol/mol reduction in HbA1c is difficult to judge at an individual level due to limited available data in the non-diabetic range, observational clinical data suggest a linear association between HbA1c and cardiovascular disease, even in non- or prediabetic individuals (*27*). For example, as a reference point, after adjusting for major conventional cardiovascular risk factors, individuals having HbA1c levels of 5.5% to 5.7% (which roughly translates to 37 to 39 mmol/mol) were almost twice as likely to be diagnosed with coronary artery disease compared to individuals with less than 5.5% (*28*). Thus, it is likely that a reduction of 1.50 mmol/mol is meaningful at the population level.

The intention-to-treat effects of the threshold rule and the causal effects of intensive lifestyle counseling for all outcomes are displayed alongside optimal bandwidth and sample size in Table 2. To ensure that the finding of improved glycemic control is not sensitive to our selected bandwidth or functional form (linear, quadratic, or polynomial), we show that effect sizes were robust to different choices (Fig. 2, table S5-6).

**Fig. 2.**
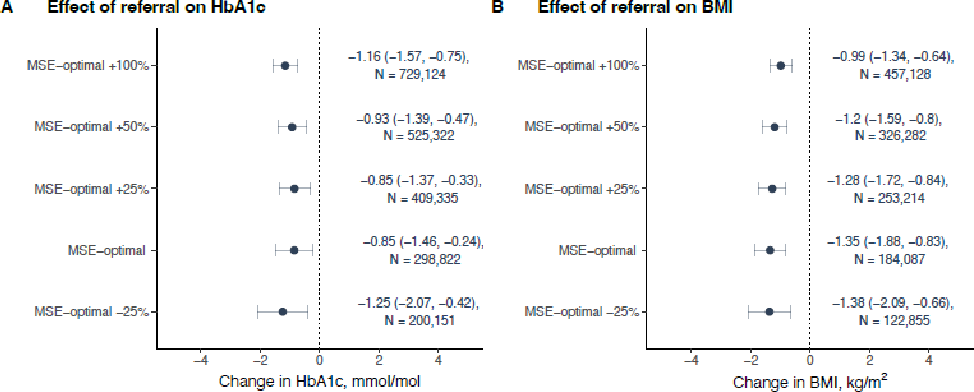
Robustness of the effects of being referred to intensive lifestyle counseling on HbA1c and BMI across bandwidth choices. MSE = mean-squared error. The mean-squared error (MSE)-optimal bandwidth is 3.8 mmol/mol below and above the threshold. The figure displays the estimated causal average complier effect of referral to intensive lifestyle counseling on (**A**) change in HbA1c and (**B**) change in BMI from local linear regressions with varying bandwidths (i.e., 75%, 125%, 150%, or 200%) of the MSE-optimal bandwidth with heteroskedasticity-robust 95% CI and triangular kernel weights. The sample size of patients in each bandwidth is given alongside the effect estimates. All effect estimates are statistically significant (p<0.05).

**Table 2.** Effect of being eligible for, and effect of being referred to, intensive lifestyle counseling on Primary and Secondary Outcomes. BMI = Body mass index. BP = Blood pressure. MACE = Major adverse cardiovascular event. MSE = mean-squared error. RD = Risk difference (i.e., difference in the probability of the outcome in percentage points). The effects were estimated in local linear regressions with heteroskedasticity-robust standard errors and triangular kernel weight. The definition of all outcomes is detailed in Table S1. We compared statistical significance (p < 0.05) to results using robust bias-corrected confidence intervals, which yielded the same statistical inferences except for diabetes medication, which was no longer significant (table S8). ^a^ Effects for HbA1c and diabetic complication were adjusted for diabetes medication prescription; effects for lipid levels were adjusted for lipid-lowering medication prescription; effects for diastolic and systolic blood pressure were adjusted for blood pressure-lowering medication; and effects for mortality and MACE hospitalization were adjusted for all three medication groups. All relevant medications are listed in our Open Science Framework project (see code availability statement). ^b^ Sample size within MSE-optimal bandwidth. ^c^ Sample restricted to those without prior lipid-lowering medication prescription. ^d^ Sample restricted to those without prior blood pressure-lowering medication prescription.

| Outcome | Unadjusted |  |  |  | Adjusted for medication <sup>a</sup> |  | MSE-optimal BW | Sample size <sup>b</sup> |
| --- | --- | --- | --- | --- | --- | --- | --- | --- |
|  | Effect of being eligible |  | Effect of referral |  | Effect of referral |  |  |  |
|  | Estimate (95% CI) | *P* value | Estimate (95% CI) | *P* value | Estimate (95% CI) | *P* value |  |  |
| Primary outcome |  |  |  |  |  |  |  |  |
| HbA1c, mmol/mol | -0.10<br>(-0.16, -0.03) | 0.006 | -0.85<br>(-1.46, -0.24) | 0.006 | -0.94<br>(-1.55, -0.34) | 0.002 | 3.8 | 298 822 |
| Secondary outcomes |  |  |  |  |  |  |  |  |
| BMI, kg/m<sup>2</sup> | -0.15<br>(-0.21, -0.09) | < 0.001 | -1.35<br>(-1.88, -0.83) | < 0.001 | - | - | 3.8 | 184 087 |
| Weight, kg | -0.33<br>(-0.48, -0.18) | < 0.001 | -2.99<br>(-4.38, -1.61) | < 0.001 | - | - | 3.4 | 208 111 |
| Diastolic BP, mmHg <sup>c</sup> | -0.16<br>(-0.4, 0.07) | 0.178 | -1.35<br>(-3.31, 0.61) | 0.178 | -1.54<br>(-3.44, 0.35) | 0.111 | 7.5 | 211 778 |
| Systolic BP, mmHg <sup>c</sup> | -0.24<br>(-0.6, 0.11) | 0.175 | -2.03<br>(-4.96, 0.91) | 0.176 | -2.41<br>(-5.21, 0.39) | 0.092 | 5.6 | 243 292 |
| HDL cholesterol, mmol/l <sup>d</sup> | 0<br>(0, 0.01) | 0.033 | 0.04<br>(0, 0.09) | 0.033 | 0.04<br>(0, 0.09) | 0.034 | 6.3 | 290 932 |
| LDL cholesterol, mmol/l <sup>d</sup> | 0.01<br>(-0.01, 0.04) | 0.265 | 0.11<br>(-0.08, 0.3) | 0.265 | 0.04<br>(-0.12, 0.21) | 0.596 | 5.7 | 107 790 |
| Total-cholesterol-to-HDL ratio <sup>d</sup> | 0<br>(-0.02, 0.02) | 0.741 | -0.03<br>(-0.20, 0.14) | 0.741 | -0.09<br>(-0.25, 0.08) | 0.286 | 5.4 | 178 852 |
| Triglycerides, mmol/l <sup>d</sup> | -0.04<br>(-0.06, -0.01) | 0.002 | -0.33<br>(-0.54, -0.12) | 0.002 | -0.32<br>(-0.52, -0.12) | 0.002 | 4.4 | 126 297 |
| Exploratory outcomes |  |  |  |  |  |  |  |  |
| Diabetes medication, RD | 0.30<br>(0.18, 0.41) | < 0.001 | 2.72<br>(1.66, 3.78) | < 0.001 | - | - | 3.5 | 501 241 |
| Antihypertensive medication, RD <sup>c</sup> | 0.11<br>(-0.38, 0.6) | 0.660 | 0.93<br>(-3.22, 5.09) | 0.659 | - | - | 4.5 | 390 878 |
| Lipid-lowering medication, RD <sup>d</sup> | 0.29<br>(-0.23, 0.82) | 0.271 | 2.61<br>(-2.03, 7.25) | 0.271 | - | - | 3.2 | 344 522 |
|  | Effect of being eligible |  | Effect of referral |  | Effect of referral |  |  |  |
|  | Estimate (95% CI) | *P* value | Estimate (95% CI) | *P* value | Estimate (95% CI) | *P* value |  |  |
| Exploratory outcomes |  |  |  |  |  |  |  |  |
| Diabetic complication, RD | 0.06<br>(-0.07, 0.19) | 0.344 | 0.58<br>(-0.62, 1.78) | 0.344 | 0.15<br>(-0.98, 1.28) | 0.797 | 3.3 | 493 431 |
| Mortality, RD | 0.05<br>(-0.09, 0.20) | 0.471 | 0.47<br>(-0.81, 1.76) | 0.471 | 0.67<br>(-0.62, 1.95) | 0.307 | 4.9 | 698 224 |
| Emergency MACE hospitalization, RD | -0.05<br>(-0.23, 0.12) | 0.550 | -0.48<br>(-2.05, 1.09) | 0.550 | -0.12<br>(-1.68, 1.45) | 0.883 | 7.5 | 211 778 |

### Confounding by medications is unlikely

It is unlikely that our finding of a beneficial effect of being referred to intensive lifestyle counseling on HbA1c is due to confounding by medication prescription or use. To rule out that we may falsely attribute improvements in glycemic control to intensive lifestyle counseling whereby they were in fact induced by diabetes medication, we adjusted our results for newly prescribed diabetes medication. In general, having an HbA1c level above the eligibility threshold for the NHS DPP was associated with a small increase in the probability of being prescribed diabetes medication shortly after treatment assignment (risk difference in percentage points [RD] = 0.04, 95% CI 0, 0.09), which increased to 0.3 percentage points at follow-up (Table 2). However, the discontinuity in the probability of being prescribed diabetes medication was not significant when using robust bias-correct confidence intervals for inference (table S8). There was no discontinuity in newly prescribed lipid-lowering medication (RD = 0.29, 95% CI −0.23, 0.82) or blood pressure-lowering medication (RD = 0.11, 95% CI −0.38, 0.60). Specifically, out of 26 513 patients with a baseline HbA1c between 42 and 47 mmol/mol who were referred to intensive lifestyle counseling, only 882 (3.3%) were prescribed diabetes medication during the 12 months following treatment assignment with numbers increasing with increasing HbA1c levels; these numbers are unlikely to substantially impact improvements in glycemic control. Indeed, when adjusting our results for being prescribed diabetes medication during follow-up, the estimated causal effect of intensive lifestyle counseling on glycemic control indicated a larger reduction in HbA1c, suggesting that improvements in HbA1c were not driven by increased uptake in diabetes medication (Table 2). We refrained from excluding patients who initiated medication from our analysis as this is likely to introduce bias given that the probability of being prescribed diabetes medication changed discontinuously at the threshold.

### Secondary outcomes provide additional evidence for health improvements at scale

In secondary analyses, we found evidence that other key cardiovascular risk factors improved. Results showed a significant association of intensive lifestyle counseling with reduced BMI (−1.35 kg/m^2^, 95% CI −1.88, −0.83) and reduced weight (−2.99 kg, 95% CI −4.38, −1.61). Albeit not significant, effect estimates were also in the direction of benefit for blood pressure levels (diastolic: −1.35 mmHg, 95% CI, −3.31, 0.61; systolic: −2.03 mmHg, 95% CI −4.96, 0.91). When adjusting results for the prescription of blood pressure-lowering medication, improvements in systolic blood pressure persisted and became marginally significant (p = 0.092; Table 2). Referral to intensive lifestyle counseling also significantly reduced triglyceride levels (−0.33 mmol/l, 95% CI −0.54, −0.12) and increased HDL levels (0.04 mmol/l, 95% CI 0, 0.09). There was no significant effect on other serum cholesterol levels (LDL and the total cholesterol-to-HDL ratio), and no effect on the probability of being prescribed lipid-lowering medication. Results were robust to adjusting for baseline observables and using different bandwidths (table S6-7, fig. fig S2-S12), a global polynomial approach (table S5), and alternative treatment definitions (table S8).

### Exploratory analyses suggest limited short-term downstream health effects

Diabetic complications, emergency hospitalization for major adverse cardiovascular events, and mortality were not significantly reduced by being referred to intensive lifestyle counseling in exploratory analyses (Table 2, fig. S13−15). The low incidence of adverse downstream events during our relatively short follow-up period, with 26 567 (1.3%) of patients dying and 36 567 (1.8% of those 2 037 384 linkable to HES data) having an emergency hospitalization for a major adverse cardiovascular event, resulted in low statistical power for detecting small, short-term changes in these outcomes. Further analyses using T2DM, hypertension, and hyperlipidemia incidence as outcomes are shown in the Supplemental Text. We interpreted these results cautiously since diagnoses in electronic health records may be less reliable than biochemical measures (*29*) and a stronger focus on identifying people who are at risk of diabetes is likely to initially increase the incidence of T2DM independent of health improvements (*30*). Lastly, we present a set of additional sensitivity analyses in the Supplement ruling out that potential sample selection effects due to immortal time bias or differential loss to follow-up may be responsible for our results (Supplementary Text, fig. S16−20, table S9−11).

### Men may be benefitting more than women

Results stratified by gender, age group, ethnicity, socioeconomic status (based on the Index of Multiple Deprivation which is derived from the patient’s postcode), and rural or urban practice location are presented in the Supplement (table S12−26). Stratification led to relatively small sample sizes for practices in rural locations and patients with Asian, Black, or Mixed ethnicity and in the youngest age group. Being referred to an intensive lifestyle intervention led to significant improvements in HbA1c, weight, blood pressure, and triglycerides in men, but not women (table S12−16). Both men and women significantly improved their BMI although effect estimates suggest larger improvements in men compared to women. There was no indication that a higher socioeconomic status was consistently associated with greater benefits (table S13−14).

## Discussion

In our study using routine, population-based electronic health records, we found causal evidence that referral to intensive lifestyle counseling led to improved glycemic control and reductions in BMI and weight. While prescriptions of diabetes medication increased slightly at the same time, sensitivity analyses demonstrated that observed improvements in glycemic control were not driven by medication. While we were not able to demonstrate a significant reduction in mortality or emergency hospitalization for major adverse cardiovascular events in the fairly short timeframe since program implementation, improvements in intermediary health markers that are key to progression to T2DM establish the potential of intensive lifestyle counseling to improve population health when implemented at scale.

### Our “real world” results are largely comparable to effects in clinical trials

An intervention should only be considered effective if it works in people who have been offered the intervention in a routine setting rather than generalizing from participants who receive the intervention in a controlled research setting (*31*). Thus, while previous studies have merely shown that intensive lifestyle counseling is *efficacious* in improving cardiovascular risk factors when performed in controlled research settings, we now present evidence that these health benefits can be successfully translated to and scaled up in routine care. Importantly—and not self-evidently— reductions in HbA1c, BMI, and weight in our study are comparable to effect sizes from clinical trials. Meta-analyses of controlled clinical trials studying effects on weight loss and blood pressure found effect sizes comparable to those presented in our study (*32–35*). These meta-analyses showed mixed effects on glucose outcomes, with both positive and null estimates having been reported (*32–34, 36*). In an early process evaluation (without a control group) of the NHS DPP (*24*), adults who attended at least one of 13 group-based intervention sessions had an HbA1c reduction of 1.26 mmol/mol and a mean weight loss of 2.3 kg. Given the fact that the wider availability and uptake of behavior change programs are currently still limited, our study lends support to calls for further investment in behavioral, population-based interventions and targeted strategies for individuals at risk for diabetes that are currently not reached through care pathways. Cardiovascular risk factors may be managed more effectively by integrating health professionals who are trained in the delivery of behavior change into primary health care teams and by improving workflow and referral processes, rather than relying on limited lifestyle advice by GPs (*37*).

Our results further suggest a decrease in the probability of being prescribed lipid-lowering medication, which appeared to have contributed to an increase, and thus adverse, effect on serum LDL at the eligibility threshold. A potential explanation may be that GPs are hesitant to concurrently refer patients to a behavior change program and prescribe lipid-lowering medication, presuming positive effects of lifestyle changes on blood lipids. At the same time, evidence from clinical trials suggests that lifestyle changes are more likely to consistently improve HDL and triglycerides levels while LDL is less likely to be affected (*35*). However, the evidence is not conclusive (*38*). Potential unintentional downstream effects of disease-specific behavior change programs, such as reduced prescriptions of medications that are likely beneficial independent of behavior change, warrant further research attention.

### Applying rigorous quasi-experimental methods to routine data can advance our understanding of health service interventions

We demonstrated the feasibility of using a regression discontinuity approach for evaluating a population-wide health service intervention by leveraging a threshold in treatment assignment induced by clinical guidelines. Thresholds are ubiquitous in clinical medicine and, thus, represent vast opportunities to generate causal, rather than associational, evidence of treatment effectiveness. In conjunction with increasing access to routine electronic health records and detailed health information, regression discontinuity analyses have the potential to greatly contribute to advancing evidence-based health care and implementation science for several reasons. First, many conceivable research questions that are of interest to clinical medicine and health systems research cannot be studied in conventional or pragmatic randomized trials, either for feasibility (such as very long follow-up periods, for example, to establish the effectiveness of anti-aging agents) or ethical reasons (such as potential harmful side effects of medical treatments) (*39, 40*). Second, since later-stage translation research questions required for population impact are frequently underfunded and understudied (*41*), applying quasi-experimental methods such as regression discontinuity to routine data could be a pragmatic approach to assessing the sustained benefit of programs similar to the NHS DPP. While social scientists are often concerned with the fact that effects estimated by regression discontinuity designs are not generalizable to observations further away from the threshold (*42*), this is less relevant to applications in clinical medicine because often patients close to the threshold are precisely those in whom we are most interested. Lastly, regression discontinuity designs may be used to investigate aspects of health equity, for example, heterogenous treatment effects between patient groups concerning sociodemographic characteristics and comorbidities, for which randomized controlled trials usually have too small of a sample size or an insufficiently diverse study population.

### Limitations and challenges in our study

Inherent to any regression discontinuity analysis is the limitation that we can only estimate the causal effect for those who initiate the treatment *because* they crossed the threshold. This effect may differ from the (unobserved) treatment effects for patients that would have received lifestyle counseling regardless of baseline HbA1c levels (the “always takers”), for example, because of clinical symptoms, or patients who would not have participated in any program or counseling even if eligible (the “never takers”). Additionally, effects may not be generalizable to those further away from the HbA1c threshold that defines prediabetes. In our analysis, we mainly relied on the CACE, the causal average complier effect, estimating the effect for those who were actually referred to an intervention. Given that there was a large percentage of individuals presenting above the HbA1c threshold who were not referred to any lifestyle counseling opportunity, it is important to not mistake the observed CACE effects for the population health effect of the NHS DPP. While the NHS DPP is operating at a large scale, with 100 000 referrals being offered in 2021 (*43*), there remains a substantial share of adults in England with impaired glycemic control who are presently not taking part in intensive lifestyle counseling, whether it would be because placements in the NHS DPP are insufficiently available or other barriers exists that decrease compliance in populations that are at risk of diabetes.

Another potential limitation is the violation of the exclusion restriction, whereby treatments or exposures other than intensive lifestyle counseling are affected by crossing the cutoff. While we could not precisely control for the relationship between drug dosage and secondary outcomes, we are confident that the observed health effects were not attributable to increased medication following treatment assignment as effect estimates were robust to adjusting for drug prescriptions. Further, it is likely that patients classified as high risk for progression to T2DM have been under closer observation by GPs and that GPs may have taken additional steps to mitigate the risk. While we cannot entirely preclude that the effects on clinical endpoints may have been caused by closer monitoring through the GP or self-care rather than intensive lifestyle counseling, we believe this is very unlikely because a placebo analysis using data before the NHS DPP roll-out provided no evidence that increased monitoring above the threshold (due to the already implemented guideline) would have led to the observed health improvements. Similarly, using electronic health records may have introduced a so-called informative observation scheme or informed presence due to differential loss to follow-up, leading to a biased observation of outcomes (*44*). Outcomes such as HbA1c or BMI were more likely to be observed in the follow-up period if patients had crossed the threshold. Since the intensity of healthcare utilization may be considered a marker of health, it may be that patients (in particular below the threshold) were more likely to consult their GP if they had other underlying health conditions or showed symptoms during the follow-up period, making patients with many visits systematically different from those with few and potentially contributing to the observed discontinuities in health outcomes. However, we addressed this concern by conditioning our results on the number of GP visits, which did not substantially change our results, and performing a sensitivity analysis restricting our sample to patients with regular GP visits before their treatment assignment.

Finally, we had to rely on what is recorded in electronic health records, e.g., we had no detailed information about adherence to behavior change programs and lifestyle counseling. However, this is only of minor concern as we are most interested in whether the positive effects of such a program would persist in the “real world”, precisely despite non-adherence to the program.

## Conclusion

The initially described skepticism about the effectiveness of lifestyle counseling for successful behavior change may stem from clinicians’ experience that brief lifestyle counseling—that is often the only feasible approach in GP consultations with pressing time constraints—may be of no or very limited effectiveness. However, we show that referrals for intensive lifestyle counseling in routine care, in the form of a population-wide diabetes prevention program, appears to be effective. Ultimately, investments in structured, intensive behavior change programs may not only reduce the risk of complications from diabetes and cardiovascular events, but their positive effects may also extend to other non-communicable diseases such as cancer, which is increasingly thought to be connected to unhealthy lifestyle habits and environments (*45*), or communicable diseases such as influenza or COVID−19, which more gravely affect people with known cardiovascular risk factors such as diabetes (*46*). Thus, our study not only demonstrates that intensive lifestyle counseling targeted at pre-diabetes can achieve important health improvements in routine care but also shows a potential route for improving population health more broadly.

## Data Availability

The data that support the findings of this study are available from Clinical Practice Research Datalink, but restrictions apply to the availability of these data, which were used under license for the current study, and so are not publicly available. Due to Clinical Practice Research Datalink license restrictions, we are unable to share data.

## Methods

### Description of the English Diabetes Prevention Programme

UK NICE public health guideline 38 recommends that people who have non-diabetic hyperglycaemia and are thus at high risk of progression to type 2 diabetes (HbA1c level of 42–47 mmol/mol [6.0–6.4%] or fasting plasma glucose of 5.5–6.9 mmol/l) are referred to a “local, evidence-based, quality-assured intensive lifestyle change programme” to prevent or delay the onset of T2DM (*1*). The NHS DPP began phased roll-out in 2016. The provider contracts require the intervention to be delivered in face-to-face to groups of 15−20 adults over at least 13 sessions (totalling 16 hours) with a minimum of 9 months’ duration, with the aim of supporting behaviour change to result in improved diet, increased physical activity and weight loss. Activities include a mixture of education, group support, knowledge testing, visual activities, and activities led by patients (*2, 3*). Individuals were identified for inclusion in the NHS DPP following an NHS Health Check, through retrospective searches of general practice records, or through routine clinical practice (*4*). According to guidelines, people below the threshold should be offered brief advice or intervention and receive information about services that could help them change their lifestyle, bearing in mind their risk profile.

### Data source and study population

The study used data from the Clinical Practice Research Datalink (CPRD) Aurum (*5*) and the NHS England Hospital Episode Statistics Admitted Patient Care (HES APC) (*6*). CPRD Aurum is a large primary care database of de-identified electronic health records from a network of General Practitioner (GP) practices across the UK. The data are representative of the broader English population in terms of geographical spread, deprivation, age, and gender (*5*). In July 2020, anonymized longitudinal data from 35.9 million patients and 1 296 currently contributing English GP practices were available. HES APC is a secondary care database in England and records patient data related to presentations in NHS hospitals or private healthcare institutions where the NHS provides partial funding.

In contrast to CPRD Aurum data, which does not contain lifetime follow-up of patients since patients are entering the database when they register with a contributing GP practice and exiting the database when they leave that practice, patients in HES APC maintain the same ID throughout their time in the database (*7*). As this information was only available for patients linkable to HES APC, we kept all patients in the analysis regardless of whether they appeared under multiple CPRD IDs. Results were insensitive to dropping all patients with multiple CPRD IDs for a single HES ID from the study population.

To ensure sufficient implementation of the NHS DPP during the study period after the phased roll-out in 2016, the population of interest consists of adults (aged 18 to 80 years) who received an HbA1c test between January 1^st^, 2017, and December 31^st^, 2018. Data were available until end of June 2020. We followed a target trial approach to mimic a randomised controlled trial that would be ideally conducted to estimate the causal program impact as closely as possible (table S1)^82,83^. Exclusion criteria were all patients who (i) exceeded the HbA1c threshold for diabetes or prediabetes prior to their index date (i.e., date of their baseline HbA1c record), or (ii) received any diabetes medication prior to their index date (all codes available in our OSF repository). Albeit specified in the NICE guideline as an alternative entry requirement, we did not use fasting plasma glucose as treatment assignment variable since routine testing of fasting plasma glucose to determine non-diabetic hyperglycemia is much less frequent as compared to HbA1c testing (*8*).

### Outcome and treatment variables

The primary outcome was glycemic control measured as change in HbA1c between baseline and the final HbA1c taken during follow-up. Secondary outcomes included change in BMI, body weight, systolic and diastolic blood pressure, serum cholesterol levels (HDL cholesterol, LDL cholesterol, and ratio between total and HDL cholesterol), and serum triglycerides level. We conducted exploratory analyses investigating the effect of program entry onto probability of newly prescribed diabetes medications, blood pressure-lowering, and/or lipid-lowering medication (evaluated separately), probability of any diabetic complication (ophthalmic, neurological, or renal), all-cause mortality, and emergency hospitalization for major adverse cardiovascular event (MACE (*9*)) during follow-up. The follow-up started at six months and continued until the date of an outcome or censoring event (such as death or transfer-out of the patient). Details on how each outcome was defined is shown in table S1.

Receipt of treatment was captured as the record of a referral to a behavior change program or intensive lifestyle counseling during the 12 months after the baseline HbA1c test. Treatment primarily included referrals to the NHS DPP, but we also included referrals to other structured programs and intensive lifestyle counseling as they are likely to serve as an alternative where placement in NHS DPP is not possible. All records considered as treatments are listed in table S4.

### Statistical analysis: Main analysis

We used a regression discontinuity analysis to estimate the association of intensive lifestyle counseling with different outcomes quantifying patients’ health status. The analysis consists of two steps: First, we estimated the association between individual’s baseline HbA1c and being referred to intensive lifestyle counseling (“first stage”). Second, we estimated the association between baseline HbA1c and each outcome by fitting separate regression lines above and belove the HbA1c eligibility threshold. The difference in where these lines intersect the threshold quantifies the discontinuity in the outcome, our effect of interest.

Specifically, our analysis represents a fuzzy regression discontinuity (FRD) design, where the treatment is not assigned deterministically but probabilistically. In the FRD design, we can estimate the intent-to-treat effect ITT_FRD_, that is, the effect of the patient presenting just above the eligibility threshold. ITT_FRD_ measures the effect of treatment eligibility, as determined by the threshold rule, and is often of interest in its own right. In particular, ITT_FRD_ can be interpreted as the effect of lowering the threshold on outcomes for the full population of patients close to the threshold. To obtain the effect of intensive lifestyle counseling itself on those induced to take up the intervention because of the threshold rule (so-called compliers), it is necessary to scale ITT_FRD_ by the difference in the probability of treatment at the threshold. This results in the complier average causal effect (CACE_FRD_).

We used a local linear approach, which minimizes bias by limiting the study sample to a defined bandwidth around the threshold in which a linear regression can be estimated (*10*). The size of the bandwidth was automatically selected using a data-driven method that seeks to optimally balance the bias-variance tradeoff (*10*). In addition, a triangular kernel was applied, such that individuals closer to the threshold were more heavily weighted than those further away. For estimation, we used a robust variance estimator and controlled for fixed effects at the practice-level. Additional analyses were performed to assess the sensitivity of results to bandwidth size and using polynomials of varying degrees.

In regression discontinuity studies, unbiased visual presentation of the data is essential (*11*). We plot the relationship between baseline HbA1c, and referral to intensive lifestyle counseling to show the discontinuity in treatment assignment. We also provide visual evidence in support of key identifying assumptions that can be tested in the data: The first is that the density of the data should be continuous around the threshold; this would be violated if patients (or providers) could precisely manipulate the running variable baseline HbA1c. The second implication is that baseline covariates should be balanced (i.e., continuous) at the threshold. As it is in randomized controlled clinical trials, evidence of balance on baseline observables provides confidence that patients assigned to treatment and control conditions are exchangeable. We used a global polynomial approach over the entire support of the data to illustrate potential unknown relationships. We did not use the global polynomial approach for causal inference procedures as it does not deliver point estimators with good properties for the treatment effect as opposed to local lower-order regressions (*12*).

### Statistical analysis: Sensitivity analyses

In sensitivity analyses, we adjusted for variables that may be associated with outcomes to show robustness and improve precision of effect estimates (*13*). These were age, gender, and number of GP consultations during follow-up. Importantly, for the primary and secondary outcomes we included time to follow-up record, i.e., months between baseline and endline measurement, and prescription of relevant medication: for HbA1c, we adjusted results for diabetes medication prescription; for blood pressure, we adjusted for blood pressure-lowering medication prescription; and for cholesterol and triglycerides levels, we adjusted for lipid-lowering medication prescription, in particular of statins, before endline measure. Estimates for mortality and MACE hospitalization were adjusted for whether patients received any of those three medications prior to their death or first MACE hospitalization.

As a retrospective study relying on routinely collected data, we expected considerable missingness in outcomes. We assessed whether this gives rise to selection bias by evaluating if missingness in outcomes or time to follow-up, changed discontinuously at the eligibility threshold. We did further sensitivity analyses to show that results are robust when (i) limiting treatment to the NHS DPP only as opposed to all intensive lifestyle counseling treatments, (ii) restricting our sample to patients for whom we observe a follow-up period of at least 24 months, and (iii) restricting our sample to patients with at least three GP consultations in three years before their baseline HbA1c. All analyses were limited to complete cases and performed using R statistical software (version 4.1.1). Mean-squared error (MSE) optimal bandwidths were automatically selected by the *rdbwselect* command of the rdrobust package (*14*). Two-sided alpha was set at 0.05.

### Heterogenous treatment effects

We present stratified, unadjusted regression discontinuity analyses, using local linear regressions with automatically selected optimal bandwidths for each subgroup, in the Supplement. Stratifying variables used in this analysis were gender (Male vs. female), age group (18−39, 40-59, 60-80), ethnicity (Asian, black, mixed or other, white; based on HES APC dataset), socioeconomic status, and practice residency (rural vs. urban). Socioeconomic status is based on the 2015 English Index of Multiple Deprivation composite score, resulting in five quintiles ranging from 1 (= “least deprived”) to 5 (= “most deprived”) and mapped via postcode of residence for patients in English practices that have consented to participate in the linkage scheme.

### Ethics

The CPRD Independent Scientific Advisory Committee approved the study protocol (20_000052) in accordance with the Declaration of Helsinki.

## Acknowledgments

This study is based on data from the Clinical Practice Research Datalink obtained under license from the UK Medicines and Healthcare products Regulatory Agency. The data is provided by patients and collected by the NHS as part of their care and support. The interpretation and conclusions contained in this study are those of the authors alone.

## Author contributions

Conceptualization: JL, CB, AJ, JD, SV, PG

Data curation: JL, MX

Formal analysis: JL, CB, MX

Visualization: JL, PG

Funding acquisition: TB, SV, PG

Project administration: JL, TB, SV, PG

Supervision: CB, TB, SV, PG

Writing – original draft: JL, PG

Writing – review & editing: JL, CB, MX, JD, AJ, SV, TB, PG

## Additional Information

Supplementary Information is available for this paper. Correspondence and requests for materials should be addressed to Pascal Geldsetzer.

